# Effects of ambient air pollution on emergency room visits of children for acute respiratory symptoms

**DOI:** 10.1101/2020.11.17.20223701

**Authors:** Rashmi Yadav, Aditya Nagori, Aparna Mukherjee, Varinder Singh, Rakesh Lodha, Sushil Kumar Kabra, Air pollution study group

## Abstract

**Background:** Data on the relation between the increase in ambient air pollution and acute respiratory illness in children are scarce. The present study aimed to explore the association between daily ambient air pollution and daily emergency room (ER) visits due to acute respiratory symptoms in children of Delhi.

**Methods:** In this epidemiological study, the daily counts of ER visits (ERV) of children (≤15 years) having acute respiratory symptoms from 1^st^ June 2017 to 28^th^ February 2019 were obtained from two general hospitals of Delhi. Simultaneously, data on daily average concentrations of particulate matter (PM_10_ and PM_2.5_), nitrogen dioxide (NO_2_), sulphur dioxide (SO_2_), carbon monoxide (CO), and ozone (O_3_), and weather variables were provided by Delhi Pollution Control Committee from their four continuous ambient air quality monitoring stations. We used K-means clustering with time-series approach to derive pollutant-derived clusters and the study period was categorized into high, moderate and low air pollution days. The combined effect of these air pollutants on acute respiratory ERV was assessed. Multi-pollutant generalized additive models (GAM) with Poisson link function was used to estimate the 0-6 day lagged change in daily ER visits with the change in multiple pollutants levels, adjusting for weather variables, days of the week and public holidays.

**Results:** In 21 months, 132,029 children were screened at the ER of the participating hospitals. Of these 19,320 (14.6%) were eligible, and 19120 were enrolled with complete data collection. The study period could be divided into 3 pollutant-derived clusters with high (Cluster 1, 150 days), moderate (Cluster 2, 204 days) low (Cluster 3, 284 days) levels of air pollution. There was a 28.7% and 21% increase in ERV among children respectively, on high and moderate level pollution days (Cluster 1 and 2) compared to low pollution days (Cluster 3) on the same day of exposure to air pollutants. Similar results were found when the exposure to ambient air pollution of previous 1-6 days were taken into account. GAM analysis showed that the association of the acute respiratory ER visits with every 10 unit change of PM_10,_ NO_2_, O_3,_ CO and SO_2_ remained significant after adjusting for multi-pollutant and confounding variables effects. In contrast, no effect was seen for PM_2.5_. The ERVs for acute respiratory symptoms rose with increase in pollutants and the trends showed a percentage change (95% CI) 1.07% (0.32, 1.83) increase in ERVs for an increase of 10 micrograms per cubic meter of NO_2_ at previous day 1, 36.89% (12.24,66.95) for 10 milligrams per cubic meter of CO at previous day 3, and 12.77% (9.51, 16.12) for 10 micrograms per cubic meter of SO_2_ at same day while decrease of −0.18% (−0.32, - 0.03) for 10 micrograms per cubic meter of PM_10_ at same day, and −4.16 % (−5.18, −3.13) for O_3_ at previous day 3.

**Conclusion:** An increase in the daily ER visits of children for acute respiratory symptoms was seen for 1-6 days after increase in daily ambient air pollution levels in Delhi.

## 1. Introduction

Air pollution is among the leading environmental risk factors contributing to the global and regional burden of diseases in India (Khilnani & Tiwari, 2018). Delhi is one of the most air-polluted cities in the world with highest annual mean particulate matter (PM)_2°5_ levels of 209°0 μg/m^3^ reported in the year 2017 (India State-Level Disease Burden Initiative Air Pollution Collaborators, 2018). Children due to their rapidly growing lungs, developing immune system, the pattern of ventilation, and outdoor activity are highly sensitive to the adverse effects of air pollution (H. Chen & Goldberg, 2009; Z. Chen et al., 2015). Children with pre-existing chronic respiratory diseases are at potentially higher risk (L. Liu et al., 2009; Segala et al., 1998). Evidence suggests that ambient air pollutants’ mixture including particulate matter (PM)_10_, PM_2.5_, nitrogen dioxide (NO_2_), sulphur dioxide (SO_2_), carbon monoxide (CO), and ozone (O_3_) have adverse respiratory health outcomes in children. These include an increase in respiratory symptoms, decreased lung function (Qian et al., 2004), increased airway inflammation (Cassino et al., 1999), aggravated asthma and chronic lung disease (*FINAL-REPORT_AQI_.Pdf*, n.d.). Several cross-sectional studies on acute effects of ambient air pollution have been comprehensivelydone in Europe, America and some of the Asian countriesviz. China, South Korea, etc. These studies have reported a positive association between ambient air pollution exposure and increase in respiratory symptoms (Preutthipan et al., 2004), daily non-accidental mortality (Cropper et al., 1997), cardio-respiratory morbidity (Rodríguez-Villamizar et al., 2018; Tapia et al., 2019), and mortality (R. Chen et al., 2012), hospitalizations (Lu et al., 2019), and emergency room (ER) visits (Babin et al., 2007; G. Chen, Li, et al., 2017; Khan et al., 2019; Pande et al., 2002). However, studies showing adverse health effects of air pollution from different parts of India are limited (Balakrishnan et al., 2011; Jayaraman & Nidhi, 2008; Maji et al., 2018; Pande et al., 2002).

In Delhi, few studies have indicated the association of acute ambient air pollution with respiratory morbidity (Agarwal et al., 2006; Jayaraman & Nidhi, 2008; Maji et al., 2018), respiratory mortality (Maji et al., 2017; Rajarathnam et al., 2011), and emergency visits for cardio-respiratory conditions (Pande et al., 2002). However, studies on acute effects of ambient air pollution on acute respiratory emergency visits are scarce. Therefore, we planned to study the impact of daily ambient air pollution on daily ER visits of children for acute respiratory symptoms in Delhi. In this work, we presented a comprehensive analysis to estimate the combined and individual effect of pollutants on pediatric ER visits in Delhi. We took a new unsupervised data-driven, machine learning approach to categorize days into bad, moderate and good air-quality days. This categorization acts as a proxy for the overall effect of pollutants from a single day which is further assessed for an increase in ER visits. We also looked at the individual impact of pollutants after adjusting for weather and holidays in single and multi-pollutant models.

## 2. Material and methods

### 2.1 Data collection

This cross-sectional study was carried out in Delhi from 1^st^June 2017 to 28^th^ February 2019. Daily counts of ER visits to hospitals were collected from two hospitals of Delhi. All children (≤15 years) visiting the ER of the All India Institute of Medical Science (AIIMS) and Kalawati Saran Children Hospital (KSCH), were screened for acute onset (less than two weeks) of respiratory symptoms by a project staff nurse and enrolled in the study. The project staff nurses trained in data collection were posted in the ER, round the clock, 24×7 for the whole duration of the data collection period. The paediatrician on duty in the ER evaluated and managed the children. Children were included if they were: (i) residing in Delhi continuously for at least four weeks, and (ii) having acute onset (≤ 2 weeks) of respiratory symptoms as well as those with exacerbations of chronic lung disease in the last two weeks. Children who were not available because of investigations or procedures and whose parents/ guardians did not consent to participate in the study were excluded from the study. After obtaining written informed consent from parents/ caretaker of eligible children the demographics, diagnosis and clinical details were recorded by the staff nurse from the patient card in the study proforma.

During the study period, daily average air pollution data from 4 continuous ambient air quality monitoring stations (CAAQMS) located in Anand Vihar, Mandir Marg, R.K. Puram, and Punjabi Bagh were obtained. Daily 24-hourly average concentrations ofPM_10_, PM_2.5_, NO_2_, SO_2_, CO, three 8-hourly concentrations of O_3_, daily mean temperature, and relative humidity data were collected from Delhi Pollution Control Committee.

The Institutional Review Boards approved the study protocol at respective institutions [AIIMS Institute ethics committee (Ref IEC/88/09.12.2015, RP-22/2015) and KSCH (LHMC/ECHR/2016/02)]. Written informed consent was obtained from the parent/guardian of the participants.

### 2.2 Statistical Analysis

The cumulative effect of daily concentration of all the six air pollutants on the proportion of ER visits due to acute respiratory symptoms ≤two weeks amongst and target population was assessed by deriving pollutant derived clusters days. K-means clustering ‘a machine learning algorithm’ was performed on the air pollutant variables to divide the study period into higher, moderate and low air pollutant days (Jin & Han, 2010; Sinha et al., 2017). The average city values of 6 ambient air pollutants acquired from 4 CAAQMS of Delhi were used. The clusters were derived based on breakpoints for Indian Air quality index (IAIQ) in the increasing order of severity for various pollutants according to Indian National Air Quality Standards (INAQS) given by Central Pollution Control Board in 2014 (Table 1)(*FINAL-REPORT_AQI_.Pdf*, n.d.; Kesavachandran et al., 2015).

**Table 1:**
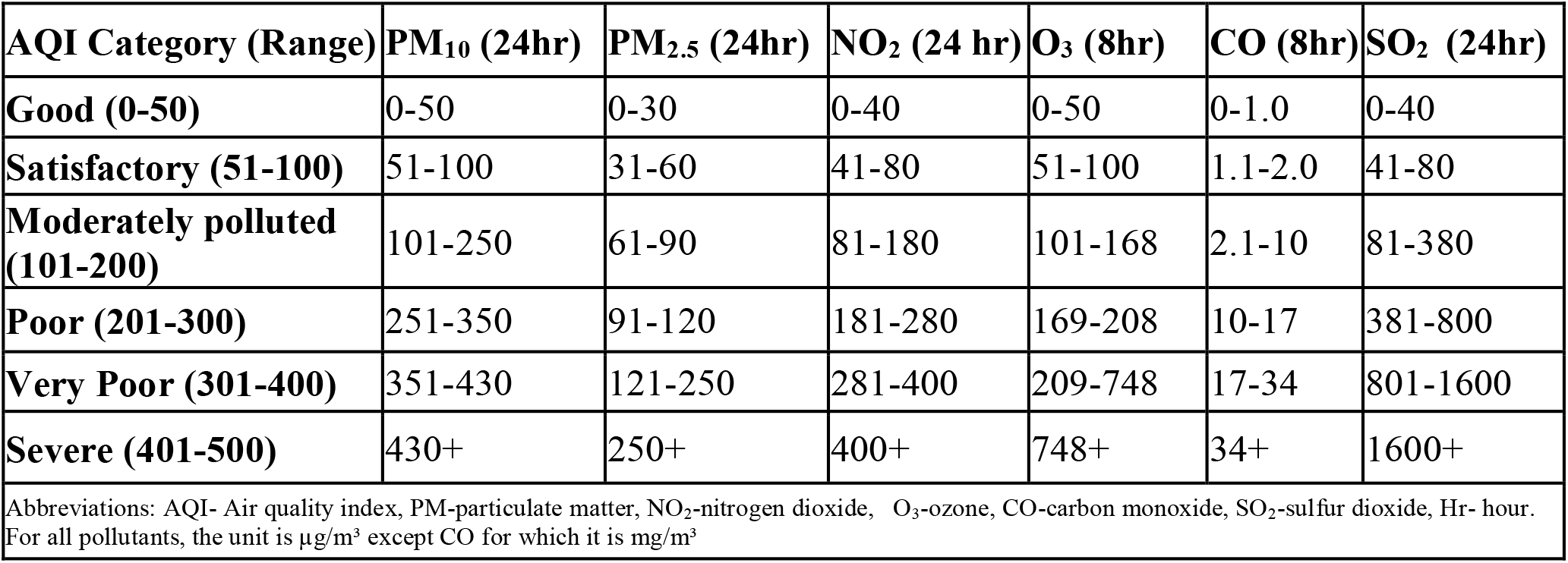
Air quality index categories and breakpoints for ambient air pollutants

The association of combined effect of ambient air pollutants levels for the same day (day 0) with the total daily ERVs and ERVs due to acute respiratory symptoms was calculated using a linear regression model. Daily percentage of enrolled children was calculated as (total daily number of enrolled children/ daily total number of children attended emergency room)*100. The percentage rise of children in high and moderate to air quality days cluster was compared to lower reference air quality day’s cluster.

Time-series analysis was done to explore the delayed association between these outcomes considering exposure to the air pollutants on previous 1-6 days. Similarly, the association between the ambient air pollutants and percentage of total daily enrolled children with individual respiratory symptoms (cough, difficulty in breathing, nasal symptoms & noisy breathing) on same days well as up to previous 1-6 days of exposure to pollutants was assessed. A generalized-additive model (GAM) with the Poisson link function was used to evaluate the association between the change in daily 24 hourly average air pollutants concentration and 1) total daily number of children attending ER, and 2) percentage total daily enrolled children with acute respiratory symptoms. Single pollutant model was run to study the individual lag effect of each of 6 air pollutant variables on the daily numbers of ER visits of children. Data was analyzed on daily timescale and accounted for the mean ambient temperature; mean relative humidity, day of the week, and public holidays. Smoothing spline function was applied to analyze the exposure-response relationship between the log-relative respiratory morbidity and the pollutants levels. The effect of pollutants was analyzed for the same day of exposure to the pollutants and considering exposure to pollutants for previous 1-3 days (Darrow et al., 2014). A multi-pollutant model was used to study the lag effect of interaction among different pollutants. The multi-pollutant model was performed for the pollutants that were significant in single pollutant analysis, and the lag day that had the highest percentage change was estimated. The data were presented as the percentage changes with 95% confidence interval (CI) in the daily acute respiratory ER visits per 10 μg/m^3^ (except 10 mg/m^3^ for CO) increase in all the pollutants concentration. All the analysis was performed using R-software version 3.6. (*R: The R Project for Statistical Computing*, n.d.).

## 3. Results

### 3.1 Descriptive statistics

During the study period, a total 1, 32,029 of children were screened who attended ER. Of these 19,320 children were eligible as they were having acute respiratory symptoms (≤2 weeks) and residing in Delhi for the past four weeks (Figure1). Of those eligible, 19120 children were enrolled.

**Figure 1:**
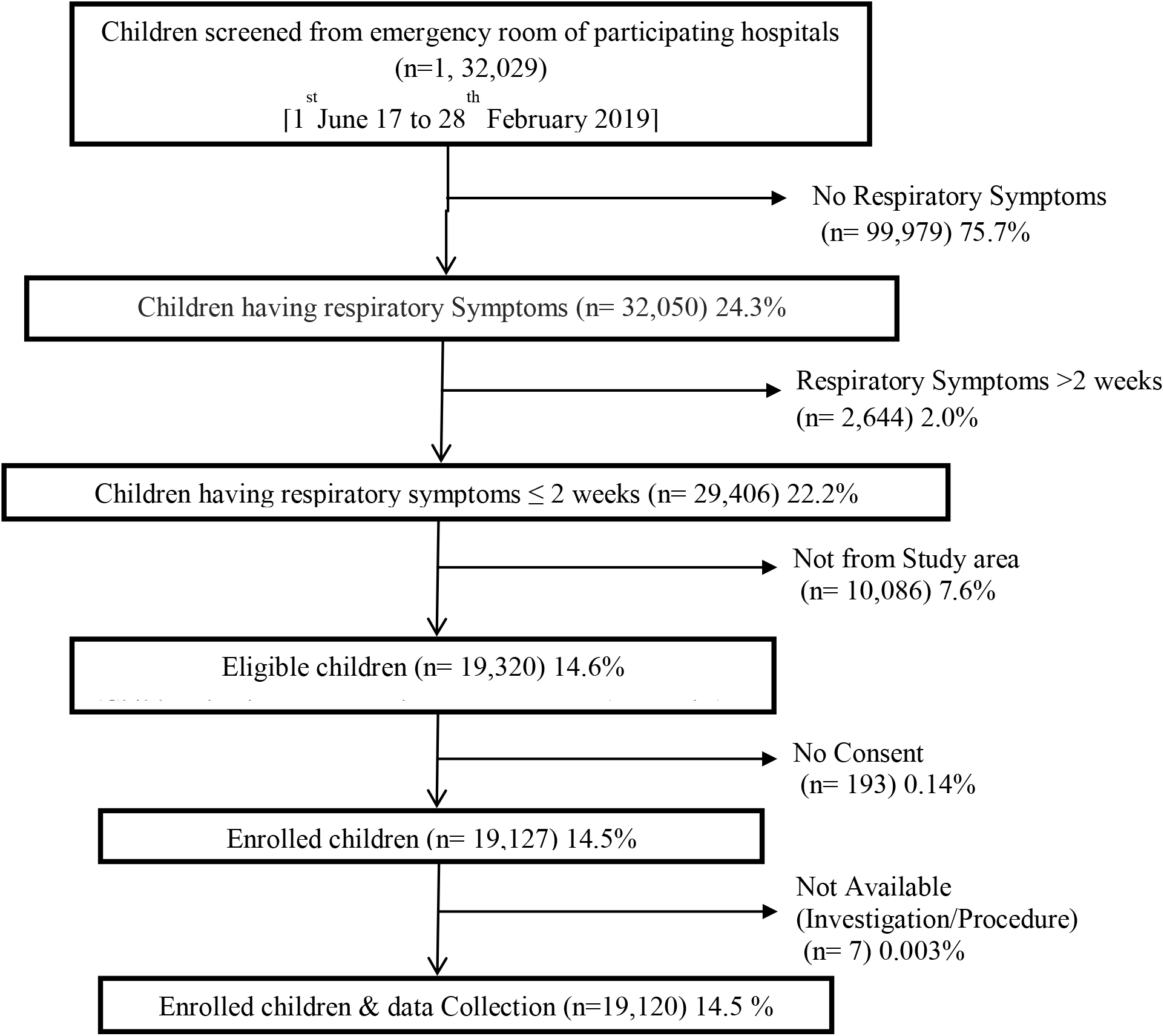
Screening and enrolment of children from emergency room of four participating hospitals during the study period

Demographic and clinical characteristics of enrolled children are shown in Table 2. The median (interquartile) age of the children was 1.1 (0.5, 3.0) years. Two-thirds of them were boys (66%). Ninety-eight per cent of children had cough, 62% had noisy breathing, 84% had difficulty in breathing, and 83% reported nasal symptoms. The average duration of acute respiratory symptoms was four days. Among the enrolled children, 85% were given ambulatory treatment, 14% were advised admission, while 1% were referred to other departments. Almost all the households were using liquefied petroleum gas for daily cooking, while approximately 0.5% used biomass or kerosene as the primary cooking fuel. The average number of household members was five withover a half living in 1-2 roomed houses. Of these, about 85% of households had a separate kitchen. Around 20% of children lived in homes where inmates regularly smoked.

**Table 2:**
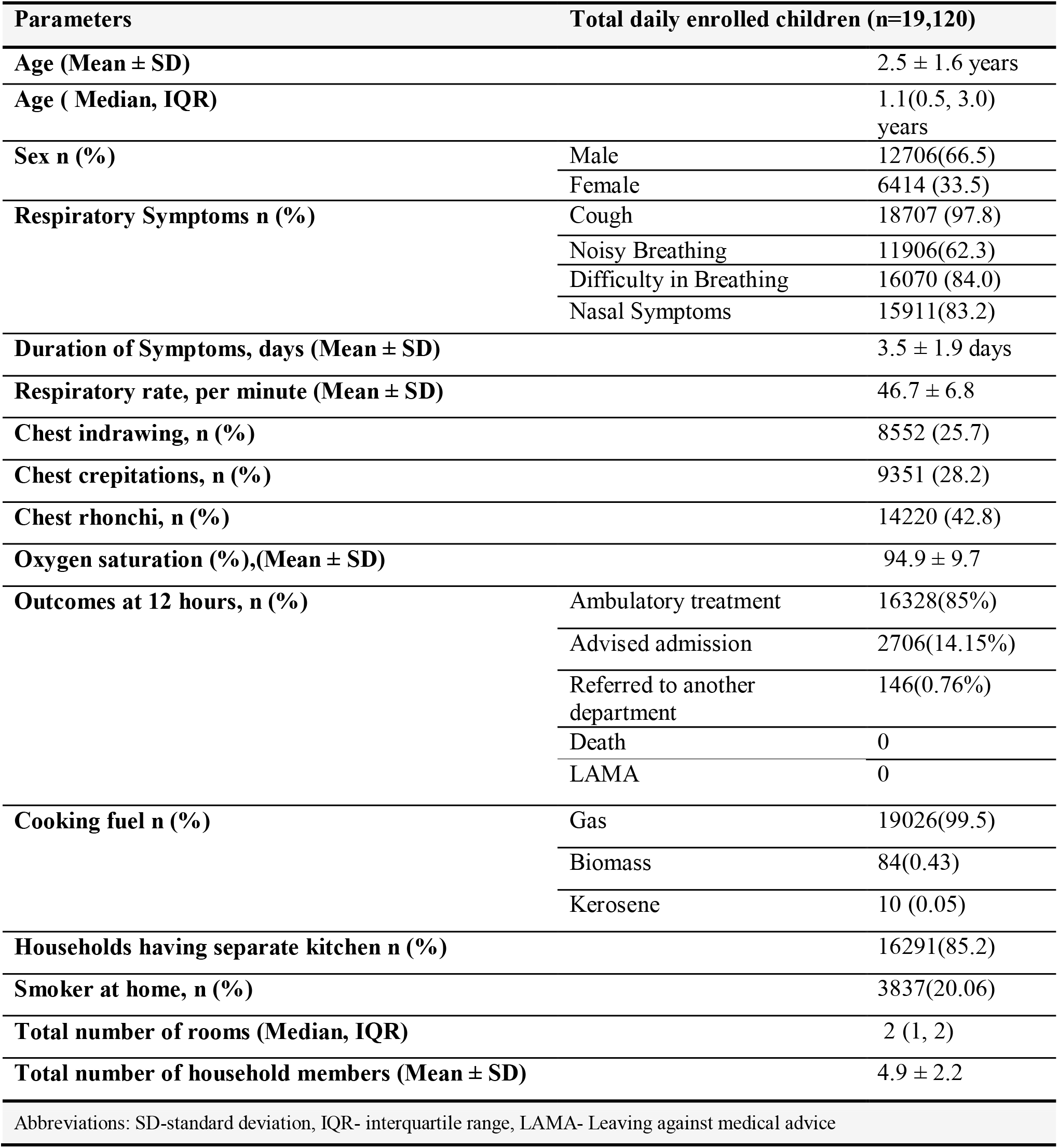
Characteristics of total daily enrolled children having acute respiratory symptoms (≤2 weeks) and residing in Delhi for the past four weeks

### 3.2 Ambient mean monthly concentration of air pollutants over the study period

The monthly trends of city average levels of six air pollutants varied during 21 months of the study period, as shown in Figure 2. From June 2017 to February 2019, two high peaks in air pollutants (a level depicting poor air quality) were observed during the months of October to December corresponding to the winter season. Another peak was found in June in the year 2018, which was possibly due to a heavy dust storm. During the summer season, (March to June) relatively low peaks of air pollutants were recorded showing moderate air quality in comparison to the winter season. The air quality was relatively satisfactory during the monsoon season from July to September.

**Figure 2:**
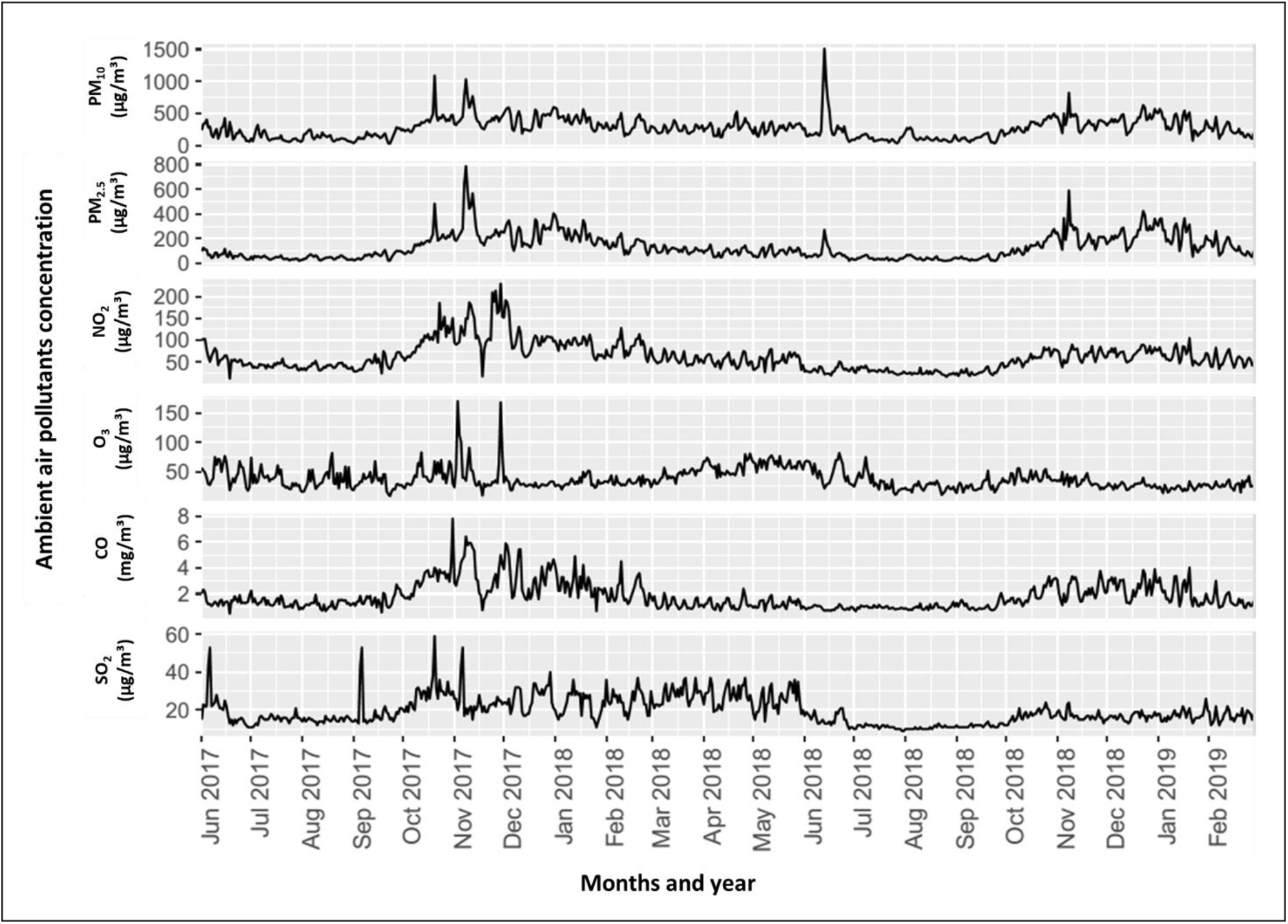
Mean monthly average levels of four pollutants during the study period (June 2017-February 2019)

### 3.3 Comparison of six pollutants levels between three clusters

The entire study duration of 674 days was divided into three clusters based on AQI based sub-categories for individual air pollutants. The three pollutant derived clusters were significantly different from each other (p<0.001 for all the pollutants), except for O_3_ (p=0.439). Figure 3 & Table 3 shows the summary statistics for six air pollutants viz. PM_10_ (µg/m^3^), PM_2.5_ (µg/m^3^), NO_2_ (µg/m^3^), O_3_ (µg/m^3^), CO (mg/m^3^), and SO_2_ (µg/m^3^) (Figure 4) in each cluster. Cluster 1 comprises of 150 days having highest levels of pollutants, e.g. median (min.-max.) levels of PM_10_ was 449 (355-1511)µg/m^3^. Cluster 2 includes 204 days with moderate levels of pollutants, e.g. median (min.-max.)levels of PM_10_ 284 (165-1018) µg/m^3^, and Cluster 3 includes 284 days with lowest levels of pollutants, e.g. median (min.-max.) levels of 134 (32-349) µg/m^3^.

**Table 3:** Descriptive statistics of daily 24 hourly levels of ambient air pollutants inthe three pollutants-derived cluster days

| Ambient air pollutants | Cluster 1<br>Higher levels of pollutants<br>(n days = 150) | Cluster 2<br>Moderate levels of pollutants<br>(n days = 204) | Cluster 3<br>Lower levels of pollutants<br>(n days = 284) |
| --- | --- | --- | --- |
| $PM_{10}$ ($\mu g/m^3$) | 449 (355- 1511) | 284 (165- 1018) | 134 (32- 349) |
| $PM_{2.5}$ ($\mu g/m^3$) | 264 (145- 787) | 134 (69- 242) | 51 (19- 91) |
| $NO_2$ ($\mu g/m^3$) | 97 (22- 230) | 62 (12- 103) | 38 (16- 82) |
| $O_3$ ($\mu g/m^3$) | 32 (14- 170) | 34 (10- 81) | 34 (9- 82) |
| CO ($mg/m^3$) | 3 (1- 8) | 1 (0.4- 3) | 1 (1- 2) |
| $SO_2$ ($\mu g/m^3$) | 22 (13- 59) | 21 (11- 37) | 14 (9- 53) |
| Data expressed as median (minimum-maximum).<br>Abbreviations: PM-particulate matter, $NO_2$-nitrogen dioxide, $O_3$-ozone, CO-carbon monoxide, $SO_2$-sulfur dioxide. | | | |

**Figure 3:**
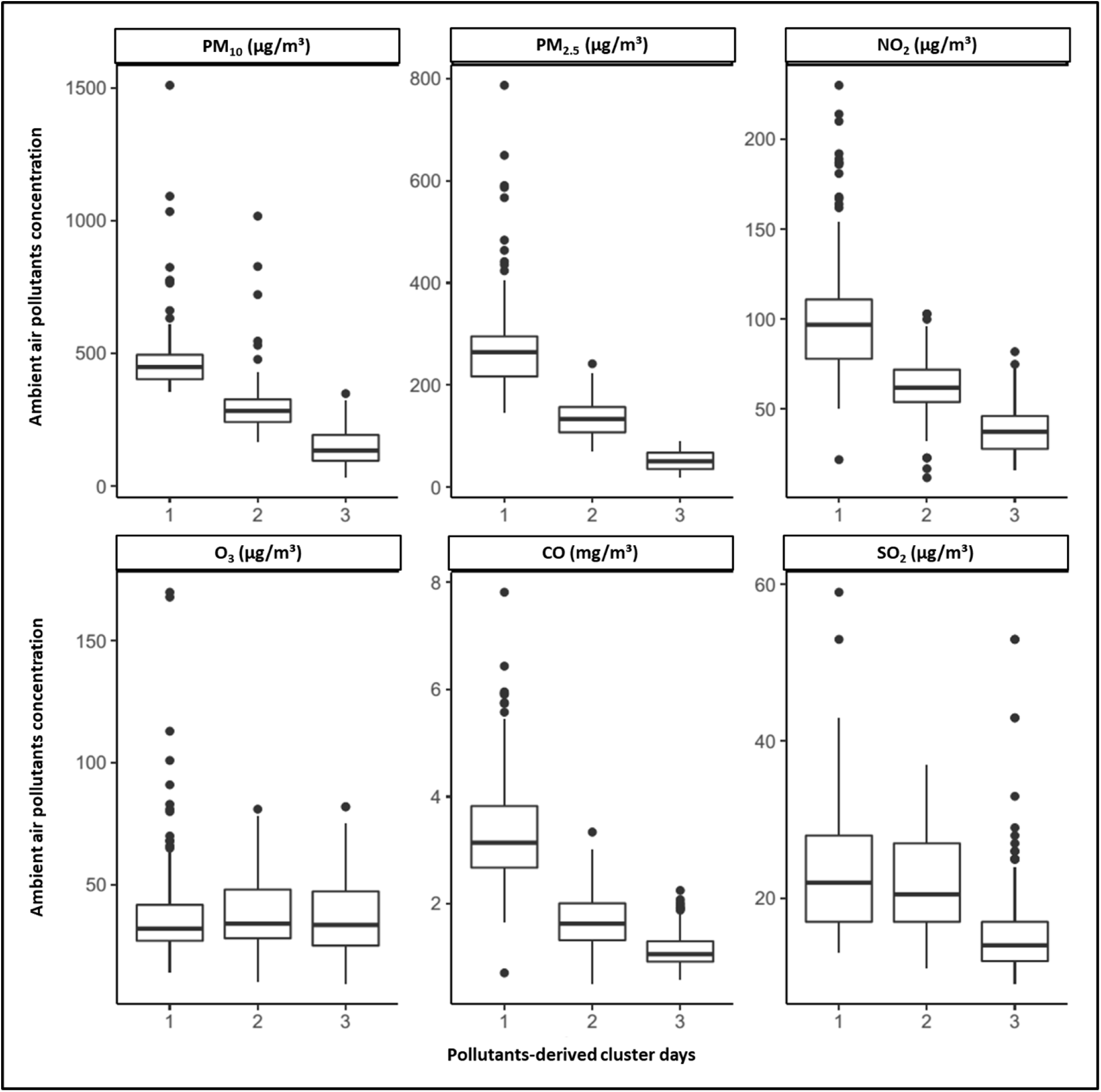
Box plot showing a comparison of each pollutant between the three pollutant-derived cluster days. Data are presented as box and whisker plots showing the median and interquartile range for air pollutants concentration. Pollutant-derived cluster days 1: higher levels of pollutants, cluster days 2: moderate levels of pollutants and cluster days 3: lower levels of pollutants.

**Figure 4:**
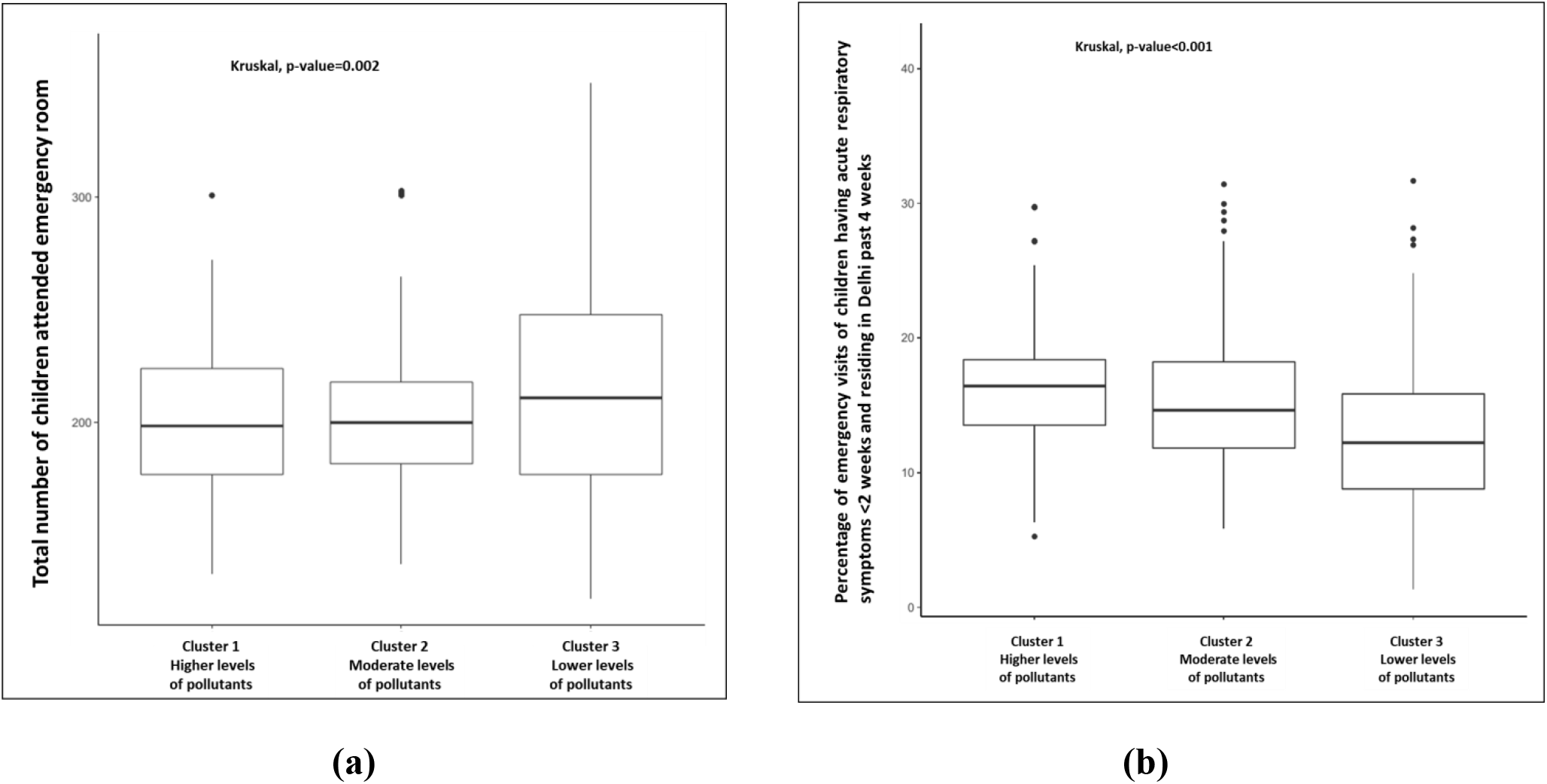
Comparison of (a) the total daily ERVs, and (b) daily ERVs due to acute respiratory symptoms between the three pollutant-derived cluster days. Data are presented as box and whisker plots showing median and interquartile range. Pollutant-derived cluster days presented as cluster 1: higher levels of pollutants, cluster 2: moderate levels of pollutants and cluster 3: lower levels of pollutants.

### 3.4 Association between ambient air pollutants and emergency room visits

#### 3.4.1 Association between air pollution and daily total ERVs and ERVs due to acute respiratory illness among children

Highest number of Total ER visits were recorded during low air pollution days in comparison to moderate and high pollution cluster days (Figure 4 a). On comparing the percentage of total enrolled children (representing children with acute respiratory symptoms <2 weeks among all children visiting ER and residing in Delhi), a significant difference was observed between the three clusters on the same day of exposure to the air pollution. There were a higher percentage of children in cluster 1 in comparison to cluster 2 & 3 (Figure 4 b).

On the day of exposure, there was 28.73 % increase in the proportion of children, having acute respiratory symptoms, enrolled daily from ER in cluster 1 and 20.96% in cluster 2 as compared to cluster 3 (Figure 5). Similar results were found when the trend of ERVs after exposure to ambient air pollution of previous 1-6 days was taken into account.

**Figure 5:**
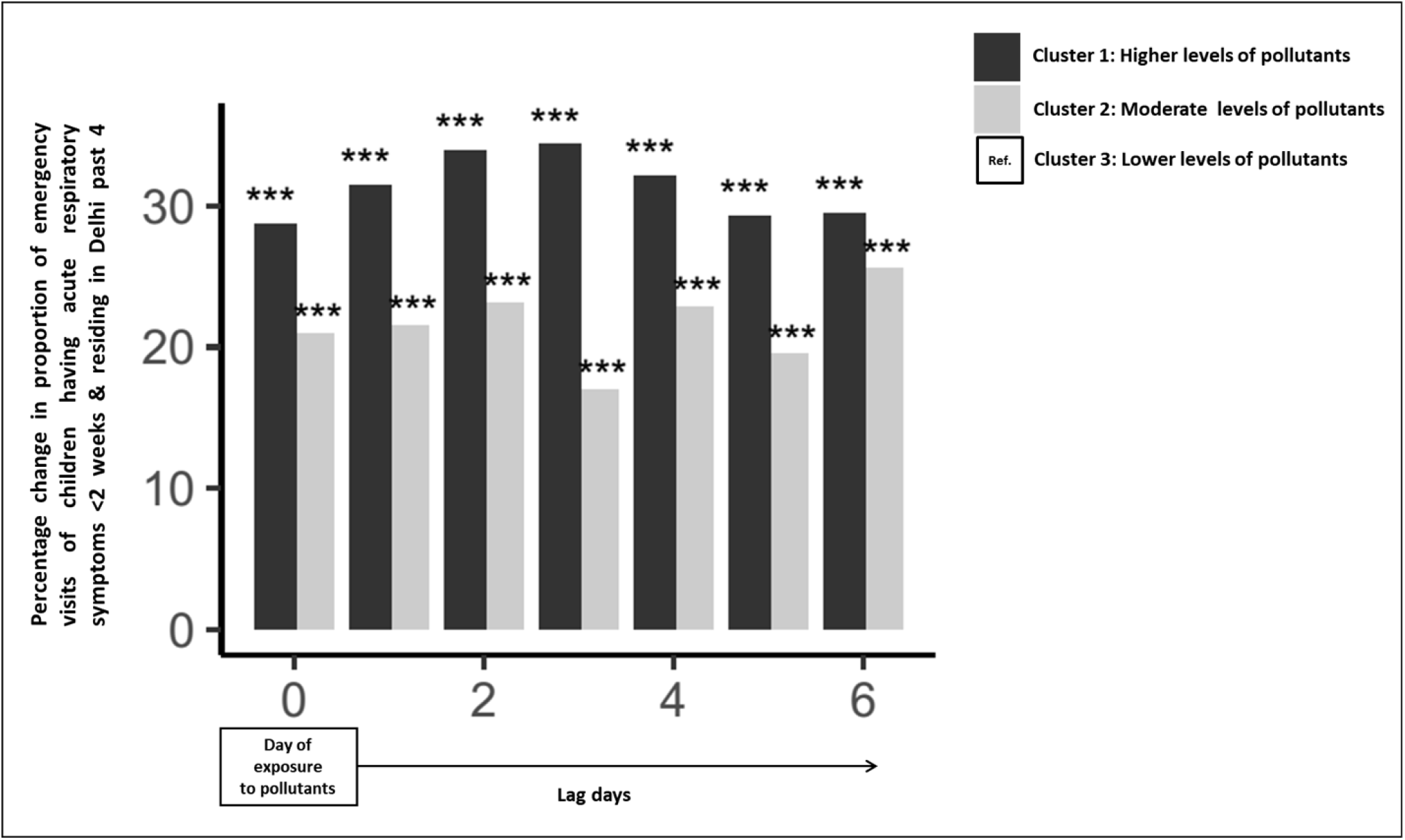
Percent increase in the proportion of total daily enrolled children attending ER with increasing air pollution [as compared with data from cluster 1]. ***p<0.0005

#### 3.4.2 Percent increase in the proportion of total dailyenrolled children with individual respiratory symptoms with increasing air pollution

The proportion of the total daily enrolled children for individual respiratory symptoms that we recorded was compared, and the percentage change was quantified. On the day of exposure, the proportion of children enrolled with cough (Figure 6A), nasal symptoms (Figure 6B), noisy breathing (Figure 6C) and difficulty in breathing (Figure 6D) was high in both cluster 1 and cluster 2 as compared to the proportion of children enrolled in the cluster 3. A similar effect was noticed when the exposure to ambient air pollution of previous 1-6 days was taken into account.

**Figure 6:**
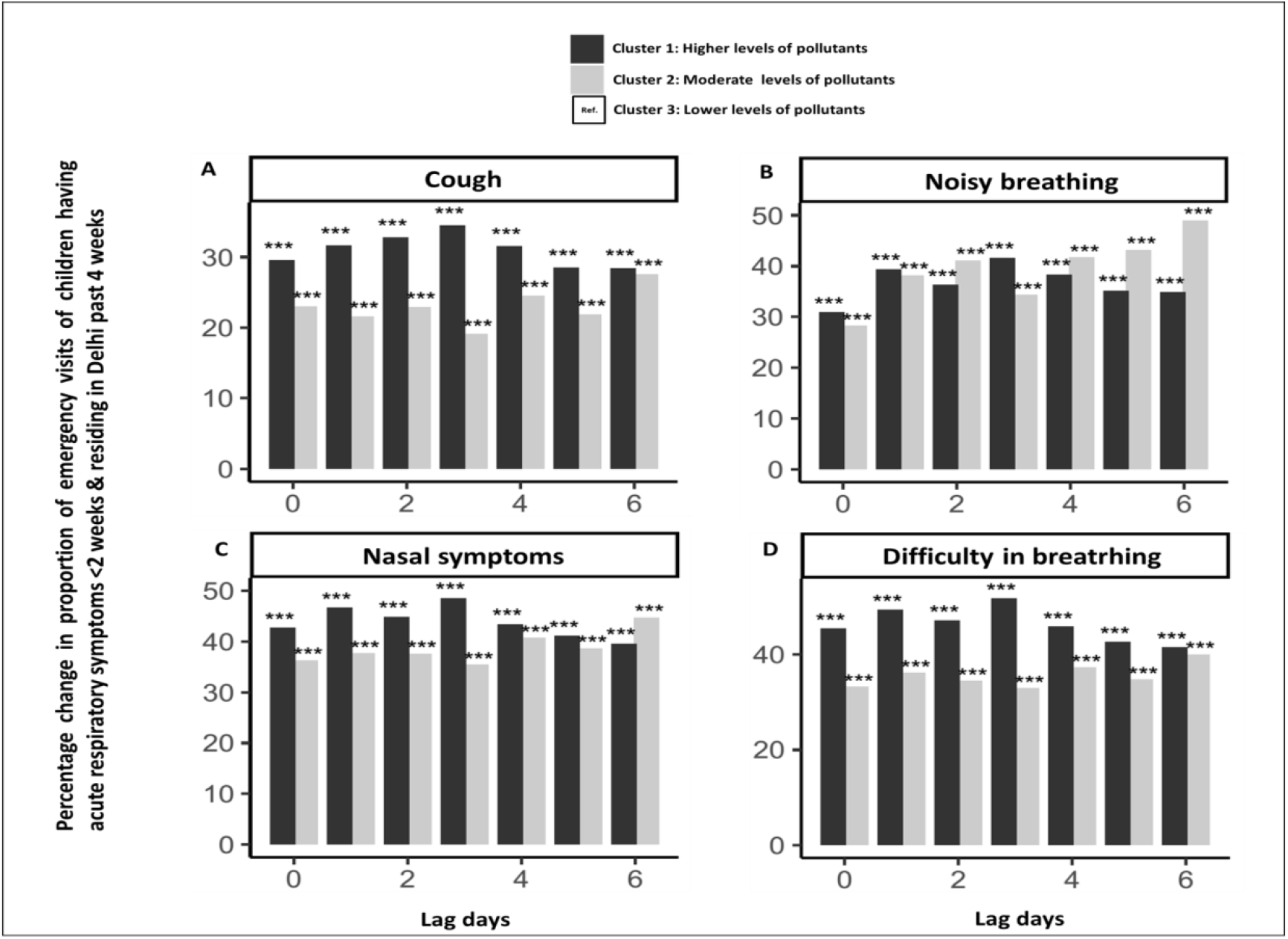
Comparison of percentage increase of the proportion of total daily enrolled children with individual respiratory symptoms for <2 weeks and residing in Delhi between the three pollutant-derived days clusters: A) Cough, B) nasal symptoms, C) noisy breathing, and D) difficulty in breathing. *p<0.05, **p<0.005, ***p<0.0005.

#### 3.4.3 Effect of individual pollutants using a multi-pollutant generalized additive model onthe total daily number of children attended ER and children with acute respiratory symptoms visiting ER

Results of the multi-pollutant models are shown in Table 4. Only the highest percentage change in ER visits of children identified from the single-pollutant model (supplementary Table 1) at a single lag day are presented. As shown in Table 4, the total daily number of children attended ER were found positively associated with PM_10,_ SO_2_ on the same day and CO on previous day 2 of exposure to pollutants, while negatively associated with PM_2.5_ on the same day and O_3_ on previous day 3 of exposure to the pollutant. The association between total daily numbers of children visiting ER having acute respiratory symptoms with the change of PM_10,_ NO_2,_ O_3,_ CO and SO_2_ was significant even after adjustment for multi-pollutant effects and confounders. However, the effect of PM_2.5_ could not be seen. Individual pollutants were found to be associated with ERvisits of children having individual respiratory symptoms such as cough, noisy breathing, and difficulty in breathing and nasal symptoms.

**Table 4.** Percentage change in daily emergency visits of children per 10-unit increase of pollutants at different lag days in the multi-pollutant model.

| Total daily number of children | Pollutant | Lag days | Single pollutant model*<br>PC % (95% CI) | Multi-pollutant model#<br>PC % (95% CI) |
| --- | --- | --- | --- | --- |
| **Attended emergency room** | PM<sub>10</sub> (µg/m<sup>3</sup>) | 0 | 0.18 (0.12, 0.25) | 0.42 (0.31, 0.52) |
|  | PM<sub>2.5</sub> (µg/m<sup>3</sup>) | 0 | 0.15 (0.02, 0.28) | -0.77 (-0.97, -0.57) |
|  | NO<sub>2</sub> (µg/m<sup>3</sup>) | 0 | 0.46 (0.05, 0.87) | - |
|  | O<sub>3</sub> (µg/m<sup>3</sup>) | 3 | -0.65 (-1.11, 0.19) | -0.92 (-1.31, -0.53) |
|  | CO (mg/m<sup>3</sup>) | 2 | 13.19 (0.10, 27.99) | 18.35 (9.01, 28.48) |
|  | SO<sub>2</sub> (µg/m<sup>3</sup>) | 0 | 5.11 (3.73, 6.51) | 5.17 (3.94, 6.43) |
| **Visited emergency room having acute respiratory symptoms** | PM<sub>10</sub> (µg/m<sup>3</sup>) | 0 | 0.33 (0.16, 0.51) | -0.18 (-0.32, -0.03) |
|  | NO<sub>2</sub> (µg/m<sup>3</sup>) | 1 | 1.31 (0.12, 2.51) | 1.07 (0.32, 1.83) |
|  | O<sub>3</sub> (µg/m<sup>3</sup>) | 3 | -1.89 (-3.11, -0.65) | -4.16 (-5.18, -3.13) |
|  | CO (mg/m<sup>3</sup>) | 3 | 29.59 (0.84, 66.54) | 36.89 (12.24, 66.95) |
|  | SO<sub>2</sub> (µg/m<sup>3</sup>) | 0 | 10.45 (6.84, 14.18) | 12.77 (9.51, 16.12) |
| **Visited emergency room having cough** | PM<sub>10</sub> (µg/m<sup>3</sup>) | 0 | 0.32 (0.14, 0.50) | - |
|  | NO<sub>2</sub> (µg/m<sup>3</sup>) | 1 | 1.9 (0.7, 3.11) | 2.10 (1.04, 3.16) |
|  | O<sub>3</sub> (µg/m<sup>3</sup>) | 2 | -2.05 (-3.48, -0.61) | -4.97 (-6.03, -3.89) |
|  | CO (mg/m<sup>3</sup>) | 1 | 47.03 (8.28, 99.65) | - |
|  | SO<sub>2</sub> (µg/m<sup>3</sup>) | 0 | 9.08 (5.46, 12.82) | 11.99 (8.7, 15.38) |
| **Visited emergency room having noisy breathing** | PM<sub>10</sub> (µg/m<sup>3</sup>) | 2 | 0.28 (-0.01, 0.56) | - |
|  | PM<sub>2.5</sub> (µg/m<sup>3</sup>) | 3 | -0.56 (-0.98, -0.14) | - |
|  | NO<sub>2</sub> (µg/m<sup>3</sup>) | 1 | -2.38 (-3.63, -1.12) | -1.84 (-2.85, -0.82) |
|  | O<sub>3</sub> (µg/m<sup>3</sup>) | 3 | -4.40 (-6.00, -2.78) | -4.63 (-6.01, -3.22) |
|  | SO<sub>2</sub> (µg/m<sup>3</sup>) | 1 | 6.90 (1.77, 12.28) | 11.60 (7.66, 15.68) |
| **Visited emergency room having difficulty in breathing** | PM<sub>10</sub> (µg/m<sup>3</sup>) | 0 | 0.47 (0.28, 0.66) | 0.59 (0.26, 0.91) |
|  | PM<sub>2.5</sub> (µg/m<sup>3</sup>) | 0 | 0.54 (0.19, 0.89) | -1.12 (-1.71, 3.38) |
|  | NO<sub>2</sub> (µg/m<sup>3</sup>) | 1 | 2.33 (1.08, 3.60) | 2.27 (1.17, 3.38) |
|  | O<sub>3</sub> (µg/m<sup>3</sup>) | 3 | -3.39 (-4.73, -2.03) | -4.51 (-5.63, -3.38) |
|  | CO (mg/m<sup>3</sup>) | 1 | 47.45 (6.59, 103.96) | - |
|  | SO<sub>2</sub> (µg/m<sup>3</sup>) | 0 | 7.75 (3.89, 11.75) | 10.64 (7.16, 14.23) |
| **Visited emergency room having nasal symptoms** | PM<sub>10</sub> (µg/m<sup>3</sup>) | 0 | 0.43 (0.25, 0.62) | 0.95 (0.64, 1.27) |
|  | PM<sub>2.5</sub> (µg/m<sup>3</sup>) | 0 | 0.39 (0.04, 0.75) | -1.69 (-2.28, -1.11) |
|  | NO<sub>2</sub> (µg/m<sup>3</sup>) | 1 | 2.28 (1.01, 3.56) | 2.08 (0.95, 3.22) |
|  | O<sub>3</sub> (µg/m<sup>3</sup>) | 0 | -1.48 (-2.83, -0.11) | -3.39 (-4.6, -2.16) |
|  | CO (mg/m<sup>3</sup>) | 1 | 62.01 (16.59, 125.13) | - |
|  | SO<sub>2</sub> (µg/m<sup>3</sup>) | 0 | 6.54 (2.63, 10.60) | 10.25 (6.73, 13.88) |

Data expressed as a percentage change (95% Confidence Interval). Statistically significant results p<0.05 are presented.
\*Highest percentage change in emergency visits of children estimated from the single-pollutant model at a single lag day after accounting for the average daily temperature, relative humidity, days of the week and public holidays.

### 3.4 Discussion

The present study examined the acute effects of air pollution by measuring daily ERVs due to acute respiratory illnesses amongchildren at two general hospitals of Delhi over 21 months. Considering the cumulative effect of all the six ambient pollutants, results of cluster analysis suggest that the daily ER visits of children having acute respiratory symptoms is higher during high and moderate air pollution cluster days on the same day as compared with low pollution cluster days. The deleterious effect was found to be sustained, as there was an increase in the respiratory illness related ERVs even when the previous day1-6 of exposure to pollutants was taken into account.

Our study results concordant withthe previous studies conducted reporting a positive association between acute effects of air pollution and 1) counts of pediatric respiratory ER visits for children (Bono et al., 2016; C. A. Lin et al., 1999; Mansourian et al., 2010) and 2) increase in respiratory symptoms among asthmatic and non-asthmatic children (Preutthipan et al., 2004). Similar to our study findings, cluster analysis studies from Kanpur, India (H.-Y. Liu et al., 2013) and China (Qian et al., 2004) have shown that individuals in high air pollution exposure cluster regions are at greater risk of having respiratory symptoms and hospital visits. However, these studiesdid not consider time lags between air pollution and the incidence of respiratory morbidity, which was evaluated in the current study. We extended this analysis on a timescale, as high pollution regions might have low pollution days. Our analysis therefore, categorized days instead of the region and looked at the future association of ER visits. Children are more susceptible to adverse health effects of air pollution than adults due to immature lung growth vulnerable to inflammatory and oxidative damage (Kuo et al., 2019). Due to higher respiration rates, and outdoor physical activity, children retain a more considerable amount of air pollutants per unit body weight than adults (Bono et al., 2016). The adverse effects of acute exposure to air pollution on respiratory health in children may be due to decreases in pulmonary function, increases in airway inflammation and oxidative stress (L. Liu et al., 2009).Various time-series studies(Bono et al., 2016; G. Chen, Li, et al., 2017; G. Chen, Zhang, et al., 2017; Rodríguez-Villamizar et al., 2018; Segala et al., 1998) have shown that any effect of air pollution may be concurrent or may last for several days after exposure to pollutants suggesting the persistence of airway inflammation and oxidative stress.

The multi-pollutant generalized additive model was carried outto examine the effect of exposure to individual pollutant levels on 0-3 day before the ER visit and their impact with other pollutants on ER visits for acute respiratory symptoms. All the air pollutants, i.e. PM_10,_ NO_2,_ O_3,_ CO and SO_2_ showed independent effects except for PM_2.5_. In the present study, a 10 µg/m^3^ increase PM_10_ was positively associated with 0.33 % (0.16, 0.51) increase in acute respiratory ER visits in the single-pollutant model, which is consistent with the earlier reports (Lee et al., 2006; C. A. Lin et al., 1999; Preutthipan et al., 2004; Rodríguez-Villamizar et al., 2018). However, in the multi-pollutant model, PM_10,_ despite having higher levels, were associated negatively with acute ER visits of children. It might be possible that during high pollution days, people try to reduce their exposure levels by staying at home, wearing masks, using air purifier etc.(R. Chen et al., 2017) thus lowering the ER visit number. The other factors that affect the degree of associations of pollutants involve average concentrations of pollutants (Lu et al., 2019), heterogeneous composition of particulate matter and gaseous mixture varying with seasons, and age and gender of the exposed population (Rodríguez-Villamizar et al., 2018; Song et al., 2018). Several studies have shown that with an increase in NO_2_, CO and SO_2,_there is a significant percentage increase in ER visits for respiratory diseases in children (Sunyer et al., 1997; Lee et al., 2006; Canova et al., 2010; Rodríguez-Villamizar et al., 2018). NO_2_ is an indicator of traffic-related air pollution emitted by vehicle exhaust (R. Chen et al., 2017; Sunyer et al., 1997). In the present study, 10 µg/m^3^ increase in NO_2_ showed delayed effect and was associated with 1.07% (0.32, 1.83) increase in acute respiratory ER visits on previous day 1 of exposure to NO_2_. Similar findings have been documented in the earlier studies (Bono et al., 2016; Rodríguez-Villamizar et al., 2018) suggesting that most of the time patient try to manage symptoms at home, which causes a delay between the onset of respiratory symptoms due to pollutant and patient reaching ER (Cassino et al., 1999).

In contrast, a 10 µg/m^3^ increase in SO_2_ concentrations showed an immediate effect on the same day associated with 12.77% (9.51, 16.12) increase in acute respiratory ER visits of children, which is in agreement with several other studies (Jaakkola et al., 1991; Kumar et al., 2007; Kuo et al., 2019; Pande et al., 2002; von Mutius et al., 1995). SO_2_ is emitted by industrial sources, domestic fuel burning and vehicles (H.-Y. Liu et al., 2013) and is a well-known immediate irritant causing inflammation of respiratory tract (Mansourian et al., 2010). For every 10 mg/m^3^ increase in CO concentrations on previous day 3 of ER visit was associated with a 36.89% (12.24, 66.95) increase in respiratory ER visits of enrolled children. This result is in concordance with previous studies showing an association of CO with ER visits (Canova et al., 2010; Villeneuve et al., 2007). CO is produced from incompletely combusted products (*FINAL-REPORT_AQI_.Pdf*, n.d.) and is postulated to exert its effect by acting as a surrogate for particulate matter (M. Lin et al., 2003). In our study period, the median concentrations of SO_2_ and CO were below permissible levels as per INAQS for the majority of the days but still showed a positive association with respiratory ER visits of children. This observation is in line with that reported in western countries (Jaakkola et al., 1991; M. Lin et al., 2003; Segala et al., 1998) in children with pre-existing chronic conditions leading to an increase in respiratory symptoms and deterioration of lung function (Jaakkola et al., 1991; Mansourian et al., 2010; Segala et al., 1998). O_3_ showed an independent negative association with acute respiratory ER on the same day as reported in earlier study (Tian et al., 2018). This lower effect of O_3_ during higher O_3_ levels could be attributed to the scavenging effects of O_3_ by the production of nitric oxide by roadside traffic (Bono et al., 2016), thereby leading to the reduced effect of O_3_ than that of PM_10_ and NO_2_ (R. Chen et al., 2017). Alternatively, the individuals predisposed to O_3_ might develop respiratory symptoms at lower concentrations and reaches the hospital before O_3_ levels reach higher levels leading to the high effect of O_3_ at lower concentration (Tian et al., 2018).

There are several limitations to our study.We used air quality data from four stationary air quality monitoring stations to represent personal exposures that may induce measurement error. Data for respiratory ER visits and ambient air quality was collected for over 1.7 years, which is often small and might limit the power of the study. Therefore, to have more accurate results,it would be helpful to carry out these studies for a longer duration, including more hospitals from every part of Delhi.

The study also has important strengths. This is the first time-series study reporting the acute effects of ambient air pollution on acute respiratory symptoms in children of Delhi. Statistical analysis was performed using a time-series analysis that provides more robust estimations (Maji et al., 2018; Rodríguez-Villamizar et al., 2018). We used a multi-pollutant model to distinguish the role of multiple pollutants taking into account day to day variations, temperature, and humidity. Such multi-pollutant modelling in time-series epidemiologic studies of acute respiratory outcomes concerning ambient air pollution is limited (Tolbert et al., 2007).We studied the health effects of 6 major pollutants mixture reflecting the ambient air pollution in contrast to the studies where the effect of only particulate matter or few pollutants were seen (G. Chen, Li, et al., 2017; Khan et al., 2019, p. 2; Kumar et al., 2007, p. 2; Linares & Diaz, 2010; Pande et al., 2002; Preutthipan et al., 2004). Preliminary findings from this study emphasize that specific policies can be made and measures can be taken to handle the increased number of children with acute respiratory problems to ER during bad air quality days. An accurate prediction can be made so that diseased children can be informed about the vulnerable effects of individual air pollutants. Long-term studies are warranted to explore the particular effect of pollutants on specific disease conditions, individual variation in exposure to local air pollution and the impact of air pollution on outpatient visits of children with chronic respiratory diseases. Additional work related to plausible biological mechanisms involved in long-term exposure of air pollution in children with respiratory disease is needed.

### 3.5 Conclusion

In the present study, we observed that short-term exposure to ambient air pollution is associated with an increase in daily ER visits of children for acute respiratory symptoms.

## Data Availability

Data is available from the corresponding authors upon reasonable request.

## Abbreviations

PM: Particulate matter
NO_2_: Nitrogen dioxide
SO_2_: Sulphur dioxide
CO: Carbon monoxide
O_3_: Ozone
ER: Emergency room
AIIMS: All India Institute of Medical Science
KSCH: Kalawati Saran Children Hospital
AQI: Air quality index
INAQS: Indian National Air Quality Standards
GAM: Generalized-additive model

## Acknowledgements

This task force project has been funded by Indian Council for Medical Research. We thank Mrs. Anumita Roy Chaudhry, from Centre for Science and Environment and Dr Arvind Pandey their valuable suggestions in this study.

## Authorship contribution statement

**Rashmi Yadav:** Data curation; Investigation; Methodology; Project administration, Validation, Visualization, Roles/Writing - original draft. **Aditya Nagori:** Formal analysis, Software, Writing - review & editing. **Aparna Mukherjee:** Formal analysis, Methodology, Software, Supervision, Writing - review andediting. **Varinder Singh:** Conceptualization, Data curation, Investigation, Methodology, Project administration, Supervision, Writing - review & editing. **Geetika Yadav:** Conceptualization, Project administration, and Visualization. **Karan Madan:** Project administration, Supervision, Writing - review & editing. **Randeep Guleria:** Project administration, Supervision and Writing - review & editing. **Raj Kumar:** Methodology, Data curation, Supervision. **Parul Mrigpuri:** Methodology, Data curation. **Rohit Sarin:** Methodology, Formal analysis, Supervision, Writing - review & editing. **Jitendra Kumar Saini:** Methodology, Formal analysis, Writing - review & editing. **Kamal Singhal:** Methodology, Data curation. **Kana Ram Jat:** Methodology, Data curation. **Rakesh Lodha:** Conceptualization, Data curation, Formal analysis, Funding acquisition, Investigation, Methodology, Project administration, Supervision, Validation, Visualization and Writing - review & editing. **Rupinder Singh Dhaliwal:** Resources; Conceptualization, Funding acquisition and Visualization. **Mohan P George:** Data curation and Methodology, Investigation. **Sushil Kumar Kabra:** Conceptualization, Data curation, Formal analysis, Funding acquisition, Investigation, Methodology,Project administration, Supervision, Validation, Visualization and Writing - review & editing. **R M Pandey and M Kalaivani**: Formal analysis.

