## Supplementary Table for "Effects of ambient air pollution on emergency room visits of children for acute respiratory symptoms"

**Table1: Percentage change in daily emergency visits of children per 10-unit increase of pollutants at different lag days in single-pollutant model**

| Total daily number of children | Lag days | $PM_{10}$ (µg/m³) | $PM_{2.5}$ (µg/m³) | $NO_2$ (µg/m³) | $O_3$ (µg/m³) | CO (mg/m³) | $SO_2$ (µg/m³) |
|---|---|---|---|---|---|---|---|
| **Attended emergency room** | **0** | 0.18 (0.12, 0.25) | 0.15 (0.02, 0.28) | 0.46 (0.05, 0.87) | | | 5.11 (3.73, 6.51) |
| | **1** | - | - | - | | | |
| | **2** | - | - | - | | 13.19 (0.10, 27.99) | |
| | **3** | -0.16 (-0.23, -0.09) | -0.24 (-0.37, -0.11) | | -0.65 (-1.11, 0.19) | | -1.66 (-2.91, -0.40) |
| **Visited emergency room having acute respiratory symptoms** | **0** | 0.33 (0.16, 0.51) | - | | | | 10.45 (6.84, 14.18) |
| | **1** | - | - | 1.31 (0.12, 2.51) | | | |
| | **2** | - | - | | | | |
| | **3** | -0.28 (-0.46, -0.1) | - | | -1.89 (-3.11, -0.65) | 29.59 (0.84, 66.54) | |
| **Visited emergency room having cough** | **0** | 0.32 (0.14, 0.50) | - | | | | 9.08 (5.46, 12.82) |
| | **1** | - | - | 1.9 (0.7, 3.11) | | 47.03 (8.28, 99.65) | |
| | **2** | - | - | | -2.05 (-3.48, -0.61) | | |
| | **3** | -0.36 (-0.54, -0.18) | - | | -2.11 (-3.36, -0.86) | | |
| **Visited emergency room having noisy breathing** | **0** | 0.26 (0.03, 0.48) | - | | | | |
| | **1** | - | - | | | | 6.9 (1.77, 12.28) |
| | **2** | 0.28 (-0.01, 0.56) | - | | | | |
| | **3** | -0.49 (-0.72, -0.26) | -0.56 (-0.98, -0.14) | -2.38 (-3.63, -1.12) | -4.4 (-6.0, -2.78) | | -5.04 (-9.09, -0.82) |
| **Visited emergency room having difficulty in breathing** | **0** | 0.47 (0.28, 0.66) | 0.54 (0.19, 0.89) | | | | 7.75 (3.89, 11.75) |
| | **1** | - | - | 2.33 (1.08, 3.60) | | 47.45 (6.59, 103.96) | 5.53 (1.34, 9.90) |
| | **2** | - | | | | | |
| | **3** | -0.34 (-0.53, -0.15) | -0.43 (-0.79, -0.07) | | -3.39 (-4.73, -2.03) | | |
| **Visited emergency room having nasal symptoms** | **0** | 0.43 (0.25, 0.62) | 0.39 (0.04, 0.75) | | -1.48 (-2.83, -0.11) | | 6.54 (2.63, 10.60) |
| | **1** | - | | 2.28 (1.01, 3.56) | | 62.01 (16.59, 125.13) | 4.3 (0.07, 8.72) |
| | **2** | - | | | -2.04 (3.65, -0.41) | | |
| | **3** | -0.41 (-0.6, -0.21) | -0.45 (-0.82, -0.09) | | -3.86 (-5.24, -2.46) | | |

Data expressed as percentage change (95% Confidence Interval). Statistically significant results $p<0.05$ are presented.

*Highest percentage change in emergency visits of children after accounting for the average daily temperature, relative humidity, days of the week and public holidays.

Abbreviations: PM-particulate matter, NO2-nitrogen dioxide, O3-ozone, CO-carbon monoxide, SO2-sulfur dioxide,.
